# Using Artificial Intelligence To Label Free-Text Operative And Ultrasound Reports For Grading Pediatric Appendicitis

**DOI:** 10.1101/2023.08.30.23294850

**Authors:** Waseem Abu-Ashour, Sherif Emil, Dan Poenaru

**Author notes:** Corresponding Author: Dr. Dan Poenaru.

## Abstract

**Purpose:** Data science approaches personalizing pediatric appendicitis management are hampered by small datasets and unstructured electronic medical records (EMR). Artificial intelligence (AI) chatbots based on large language models (LLMs) can structure free-text EMR data. Here we compare data extraction quality between ChatGPT-4 and human data collectors.

**Methods:** To train AI models to grade pediatric appendicitis preoperatively, several data collectors (medical students and research assistants) extracted detailed preoperative and operative data from 2100 children operated for acute appendicitis between 2014-2021. Collectors were trained and certified for the task based on satisfactory Kappa scores. ChatGPT-4 was prompted to structure free text from 103 random anonymized ultrasound and operative records in the dataset using the set variables and coding options, and to estimate the Pediatric Appendicitis Grade (PAG) from the operative report. A pediatric surgeon then adjudicated all data, identifying errors in each method.

**Results:** Within the 44 ultrasound (42.7%) and 32 operative reports (31.1%) discordant in at least one field, 98% of the errors were found in the manual data extraction. The PAG was erroneously assigned manually in 29 patients (28.2%), and by ChatGPT-4 in 3 (2.9%). Across datasets, the use of the AI chatbot was able to avoid misclassification in 59.2% of the records including both reports and extracted data approx. 100x faster than manually.

**Conclusion:** An AI chatbot significantly outperformed manual data extraction in accuracy for ultrasound and operative reports, and correctly assigned the PAG score. While wider validation is required and data safety concerns must be fully addressed, these novel AI tools show significant promise in improving the accuracy and efficiency of research data collection.

**Highlights:** 1. What is known about this topic?

AI chatbots have several benefits and implications including healthcare uses.

2. What new information is contained in this article?

AI chatbot was proven to be more accurate when compared to human data extraction.

## Introduction

Perforated appendicitis comprises approximately 25%–30% of appendicitis cases and has historically been considered a single disease entity [1]. Current clinical scoring systems for appendicitis, such as the Pediatric Appendicitis Score and the Alvarado score, focus on diagnosis of the disease, rather than its severity [2]. Attempts have also been made to achieve a standard definition for perforated appendicitis [3]. However, a recent review of the outcomes of perforated appendicitis revealed persistent significant variability in the outcomes of perforated appendicitis owing to lack of utilization of an evidence-based definition [4]. Previous work within our own team has protocolized the care of children with perforated appendicitis by developing a complication grade [5] and has shown that postoperative outcomes and resource utilization strongly correlate with increasing grade of perforated appendicitis [3].

Artificial intelligence (AI) and its subset of machine learning (ML) have demonstrated significant potential in pediatrics and in surgery [6]. Specifically for acute appendicitis, various AI methods have been explored for improving the accuracy of acute appendicitis diagnosis and predicting the need for surgery [7,8]. Clinical prediction tools (CPTs), using both patient data and details from patient encounters, have become an integral part of diagnosis and treatment processes, especially with ML approaches [9]. ML-based CPTs have shown to be superior in silico, such as in diagnosing suspected appendicitis in pediatric populations [10], due to their capability to handle complex data like electronic medical records (EMRs). However, their clinical applicability and validation in clinical settings remain underexplored [10]. The considerable increase in health data digitization over the past decade has seen a rise in multimodal data repositories, including electronic health text data, medical imaging, multiomics, and environmental data [11]. However, deriving robust real-world evidence from these data requires overcoming barriers like data recency, clinical depth, provenance, completeness, representativeness, and usability [12]. Conventionally, extracting clinical variables and outcomes from routinely collected EMRs necessitates significant pre-processing, laborious curation, and intensive manual patient chart reviews, posing a time and resource challenge [13].

Lately, the field of AI has seen an expansion of deep learning, an emergent AI branch, into numerous network architectures. While ML typically involves the use of statistical methods to learn from data, deep learning uses neural networks to learn from large datasets [14]. Several healthcare applications of deep learning are frequent today, such as computer vision for radiology and pathology, speech recognition, and natural language processing (NLP) [15]. AI’s transformative power in healthcare is demonstrated in its ability to analyze large volumes of data, informing treatment decisions, enhancing medical research, and facilitating tasks like early disease diagnosis, outcome predictions, and automation of routine tasks [16]. Chatbots represent AI-driven applications that mimic human dialogue, offering automated responses to user queries through the use of NLP [17]. They serve various roles including education, health assistance, and financial management [18]. Well-known examples of chatbots include Siri, Alexa and Google Assistant [19]. ChatGPT, recently appeared, has proven to be a useful chatbot. This AI system utilizes ML algorithms and NLP to generate human-like responses, functioning as a sophisticated chatbot [20,21]. Despite not being originally intended for healthcare applications, it exhibits potential in supporting patient queries and easing healthcare operations [22]. The automation of tasks like reliable and swift transcription of patient medical records allows medical professionals to dedicate more time to patient interactions [23]. In making medical reports, clinical trial documents, and other related documents more comprehensible for both patients and healthcare professionals, ChatGPT can serve as a summarizing tool which can potentially reduce the risk of errors in medical records [24,25]. ChatGPT’s ability to translate medical text between languages also contributes to improved communication and understanding among patients and healthcare providers [24,25].

In this pilot study, we compared clinical data extraction from free-text medical reports between an AI chatbot, ChatGPT-4, and trained human data extractors.

## Methods

### Study Design

This is a *<u>fully paired</u>* comparative accuracy study [26] to compare the accuracy of ChatGPT-4 and human data collectors.

### Patient Characteristics

Our dataset included pediatric patients from the Montreal Children Hospital (MCH) between January 2014 - December 2021. Patient criteria for inclusion were children ≤ 18 years of age who underwent surgery for acute appendicitis and were confirmed to have acute appendicitis. Patients were excluded if they were <1 or > 18 years old at the time of operation, had incomplete medical records, or if no conclusive diagnosis or grade of perforation was possible from the operative report.

### Instruments and Data Collection

A standard case report form was created in the secure web-based Research Electronic Data Capture (REDCap) software. Patient demographics were collected via the Outcome and Assessment Information Set (Oacis) platform, the McGill University Health Centre (MUHC) electronic health record platform. To develop an appendicitis severity prediction model, we extracted data from two report types: ultrasound (US) and operative reports.

### Perforated Appendicitis Grade (PAG)

The PAG **(Table 1)** has been developed in the H.E. Beardmore Division of Pediatric Surgery based on an earlier validated appendicitis score [5]. It was used directly to assign a score of 0-5 in a selection of operative reports.

**Table 1:** Perforated Appendicitis Grade (PAG) Definition And ChatGPT-4 Prompt Definitions.

| PAG | Grade Name | Definition | Additional OR Terms | Additional US Report Terms |
| --- | --- | --- | --- | --- |
| **0** | Normal Appendix | No signs of inflammation | Normal<br>Unremarkable/ white appendix | Normal examination |
| **1** | Appendicitis Without Perforation | No visible hole in the appendix.<br>No free fecalith. No extravasation of appendiceal contents in vivo or ex vivo.<br>Gangrenous appendix. | appendix non-perforated<br>simple appendicitis<br>Appendix inflamed/phlegmonous/gangrenous/dilated/necrotic/injected/edematous/non-compressible | No |
| **2** | Early Or Contained Perforation | Visible hole or free fecalith or extravasation of appendiceal contents in vivo or ex vivo. Pus and/or fibrinopurulent exudate limited to right lower quadrant and/or pelvis. | Perforated appendicitis<br>Complex appendicitis<br>Appendix perforated/leaking pus<br>pus/fluid/collection around the appendix<br>Appendix sealed /covered by omentum | Fat stranding/echogenic fat/mesenteric fat stranding<br>Fluid collection around appendix<br>Fecalith outside the appendix<br>Phlegmon/inflammatory mass |
| **3** | Perforation With Abscess | Discrete cavity containing pus not in free communication with the peritoneal cavity. No pus or fibrinopurulent exudate outside right lower quadrant and/or pelvis, or between bowel loops. | Perforated appendicitis<br>Complex appendicitis<br>Single abscess<br>Break into / unroof a collection/<br>pocket of pus<br>Interloop abscess | Fat stranding/echogenic fat/mesenteric fat stranding<br>Fecalith outside the appendix<br>Abscess / organized fluid collection |
| **4** | Perforation With Generalized Peritonitis | No discrete abscess. Pus and/or fibrinopurulent exudate extending outside the right lower quadrant and/or pelvis to involve at least one of the following: right upper quadrant, left upper quadrant, left lower quadrant, interloop spaces. | Perforated appendicitis<br>Complex appendicitis<br>Generalized peritonitis<br>Pus in all/four quadrants<br>Diffuse peritonitis<br>Fibrinopurulent peritonitis | Fat stranding/echogenic fat/mesenteric fat stranding<br>Fecalith outside the appendix<br>Free fluid |
| **5** | Perforation With Abscess & Generalized Peritonitis | Features of Grades 3 & 4 | Perforated appendicitis<br>Complex appendicitis<br>Single or multiple abscesses<br>Break into / unroof collection(s)<br>Pocket(s) of pus<br>Interloop abscess(es)<br>Pus in four quadrants with abscess<br>Diffuse peritonitis<br>Fibrinopurulent peritonitis | Fat stranding/echogenic fat/mesenteric fat stranding<br>Fecalith outside the appendix<br>Abscess / organized fluid collection |

### Determination of perforation and severity of appendicitis (OR report)

The operative reports served as the reference standard for determining appendiceal perforation and severity of appendicitis. If there was a contradiction between the operative and pathology reports regarding the presence of perforation, the pathology report was considered definitive.

### Procedures

To train AI models to grade pediatric appendicitis preoperatively, several data collectors (medical students and research assistants) extracted detailed preoperative and operative data from 2100 children operated for acute appendicitis between January 2014 and December 2021. Human data collectors were trained to extract the necessary clinical data into our dataset and derive the PAG. For the current study, a random selection of 103 ultrasound and operative reports were tested as prompts for ChatGPT-4, in conjunction with the PAG. For this purpose, the PAG categories were expanded using an expert-derived vocabulary of pertinent medical terms and expressions extracted from actual operative reports. ChatGPT was prompted to structure the free text of both report types using set parameters representing the actual variables and codes in the dataset, and to estimate the PAG from the operative report **(Supplemental File [A])**. A pediatric surgeon then compared the human and AI-generated data, identifying all misclassified data within each group.

### Variables and statistical analysis

The patient data used for this report was part of a larger dataset on pediatric patients with appendicitis. For this study we limited information to be collected to the US and the operative report. The data collected from the *US report* included: ultrasound diagnosis, appendix identification, appendiceal maximum diameter, appendix location, probe tenderness over appendix, appendix compressible, mesenteric fat stranding, fluid around appendix, fluid in pelvis, phlegmon or inflammatory mass, bowel thickening, presence of fecalith, intra-abdominal abscess, as well as the full US report text. The data collected from the *operative report included*: appendiceal necrosis, fecalith, intra-abdominal abscess, intraperitoneal fluid, peritonitis, presence of perforation, site of perforation (if mentioned), PAG recorded in report, intraoperative PAG mentioned in the report, an automatically calculated PAG within REDCap, and the full operative report text.

All de-identified information was exported into an Excel sheet. We calculated percentages and rates of discrepancy between human data extractors and ChatGPT-4. Data was organized and preliminary analyses were conducted using Microsoft Excel (version 16.74, 2023, Microsoft Corp.). Raw data was entered into Excel spreadsheets, and data cleaning was performed to check for discrepancies between human data extractors and ChatGPT-4. We calculated percentages and rate of errors between human data extractor information and ChatGPT-4 output.

## Results

We compared the data between human data extractors and ChatGPT-4 in 103 records. The data collected included 13 variable fields from the US report and 9 fields from the operative report. The included 103 records were a subset of a larger multimodal dataset gathered for a ML appendicitis classification project. The entire dataset will contain 2300 records. Within this large dataset, missingness of information related to the US and operative reports varied widely. We ensured however that the 103 records selected for this analysis did not contain any missing information. The actual PAG distribution in these 103 records was: grade I - 24%, II - 12.5%, III - 11.5%, grade IV - 10%, and grade V - 42%. The correction made by ChatGPT-4 to human extractors across all records encompassing both types of reports was found in 61 records (59.2%).

### US Reports

We compared 13 fields extracted from the US reports by human and AI extractors (**Table 2)**. **Figure 1** shows the US report human extractors read and the structured ChatGPT-4 output after using the prompt. The discordance between human data extractors and ChatGPT-4 in the US report features ranged from 0% - 12.6%. The highest discrepancies were in the final US diagnosis (12.6%), followed by presence of intra-abdominal abscess (11.6%), mesenteric fat stranding (7.8%) and fluid in the pelvis (7.8%). There were no discrepancies encountered in appendiceal compression. Based on the ground truth assessment by the expert, ChatGPT-4 was found to have successfully corrected the human data extractors in 65 individual fields within 44 records, yielding an overall 42.7% correction rate. The misclassification rate by Cahatgpt-4 was in only 9 individual fields within (8.7%) of the records.

**Table 2:** Accuracy And Discrepancies In US Report Field By Data Extraction Method.

| **Field** | **Discrepancies % (n),**<br>**Total = 103** | **Errors in Human**<br>**Extractors % (n),**<br>**Total = 103** | **Errors in**<br>**ChatGPT-4 %**<br>**(n),**<br>**Total = 103** |
| --- | --- | --- | --- |
| *Ultrasound diagnosis* | 12.6% (13) | 9.7% (10) | 2.9% (3) |
| *Appendix identified* | 3.9% (4) | 3.9% (4) | 0.0% (0) |
| *Appendiceal max.*<br>*diameter* | 5.8% (6) | 5.8% (6) | 0.0% (0) |
| *Appendix location* | 3.9% (4) | 3.9% (4) | 0.0% (0) |
| *Probe tenderness*<br>*over appendix* | 1.0% (1) | 1.0% (1) | 0.0% (0) |
| *Appendix*<br>*compressible* | 0.0% (0) | 0.0% (0) | 0.0% (0) |
| *Mesenteric fat*<br>*stranding* | 7.8% (8) | 7.8% (8) | 0.0% (0) |
| *Fluid around*<br>*appendix* | 5.8% (6) | 4.8% (5) | 0.97% (1) |
| *Fluid in pelvis* | 7.8% (8) | 7.8% (8) | 0.0% (0) |
| *Phlegmon or*<br>*inflammatory mass* | 5.8% (6) | 5.8% (6) | 0.0% (0) |
| *Bowel thickening* | 4.8% (5) | 4.8% (5) | 0.0% (0) |
| *Presence of fecalith* | 0.97% (1) | 0.0% (0) | 0.97% (1) |
| *Intra-abdominal*<br>*abscess* | 11.6% (12) | 7.8% (8) | 3.9% (4) |
| ***ChatGPT-4 improved human extracted records = 42.7% (44)*** |  |  |  |

**Fig. 1:**
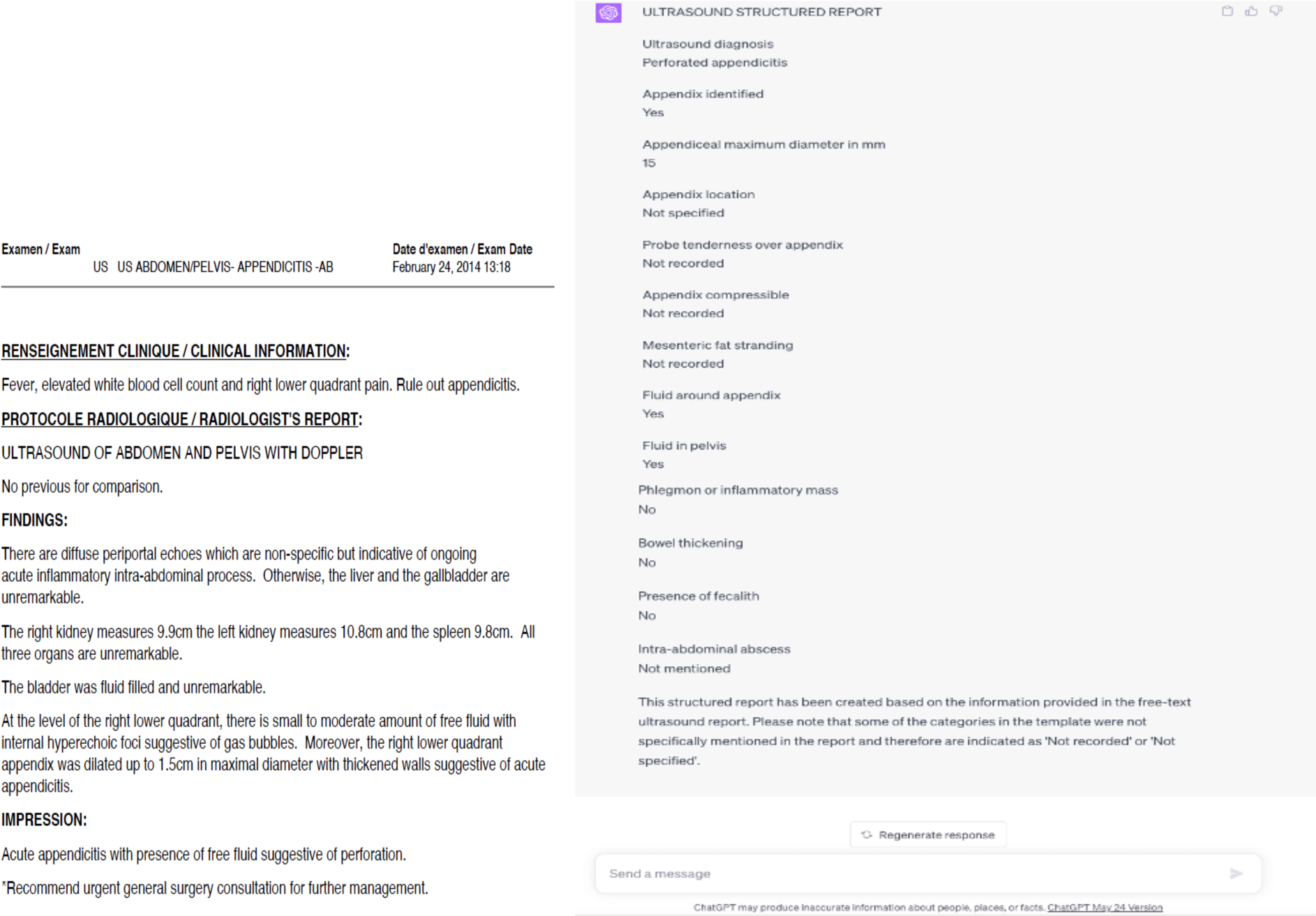
US Report & ChatGPT-4 Structured Data After Prompt.

### Operative Reports

Within the operative reports we compared 10 distinct fields (**Table 3**). **Figure 2** shows the operative report human extractors read and the structured ChatGPT-4 output after using the prompt. The discrepancy between human data extractors and the ChatGPT-4 the operative report ranged from 0% - 41.7%. The highest discrepancy was in the final PAG (31%), followed by presence of peritonitis (26.2%), intra-operative PAG (20.4%) and intra-abdominal abscess (18.4%). Following validation by the expert surgeon, ChatGPT-4 corrected human data extractors in 133 individual fields, yielding a 31.1% overall correction rate. Importantly, the PAG was misclassified by the human collectors in 28% of the records, and only in 3% by ChatGPT-4. **Figure 3** shows human vs ChatGPT-4 errors in each report.

**Table 3:** Accuracy And Discrepancies In Operative Report Field By Data Extraction Method.

| **Field** | **Discrepancies % (n),**<br>**Total = 103** | **Errors in Human**<br>**Extractors % (n),**<br>**Total = 103** | **Errors in**<br>**ChatGPT-4 %**<br>**(n),**<br>**Total = 103** |
| --- | --- | --- | --- |
| *Appendiceal necrosis* | 6.8% (7) | 6.8% (7) | 0.0% (0) |
| *Fecalith* | 5.8% (5) | 4.8% (5) | 0.0% (0) |
| *Intra-abdominal*<br>*abscess* | 18.4% (19) | 18.4% (19) | 0.0% (0) |
| *Intraperitoneal fluid* | 6.8% (7) | 6.8% (7) | 0.0% (0) |
| *Peritonitis* | 26.2% (27) | 26.2% (27) | 0.0% (0) |
| *Presence of*<br>*perforation* | 5.8% (6) | 5.8% (6) | 0.0% (0) |
| *Site of perforation*<br>*mentioned* | 0.0% (0) | 0.0% (0) | 0.0% (0) |
| *Intraoperative*<br>*perforation grade* | 20.4% (21) | 20.4% (21) | 0.0% (0) |
| *PAG* | 31.1% (32) | 28.1% (29) | 2.9% (3) |
| ***ChatGPT-4 improved human extracted PAG Grade = 28% (29)*** |  |  |  |
| ***ChatGPT-4 improved human extracted records = 31.1% (32)*** |  |  |  |

**Fig. 2:**
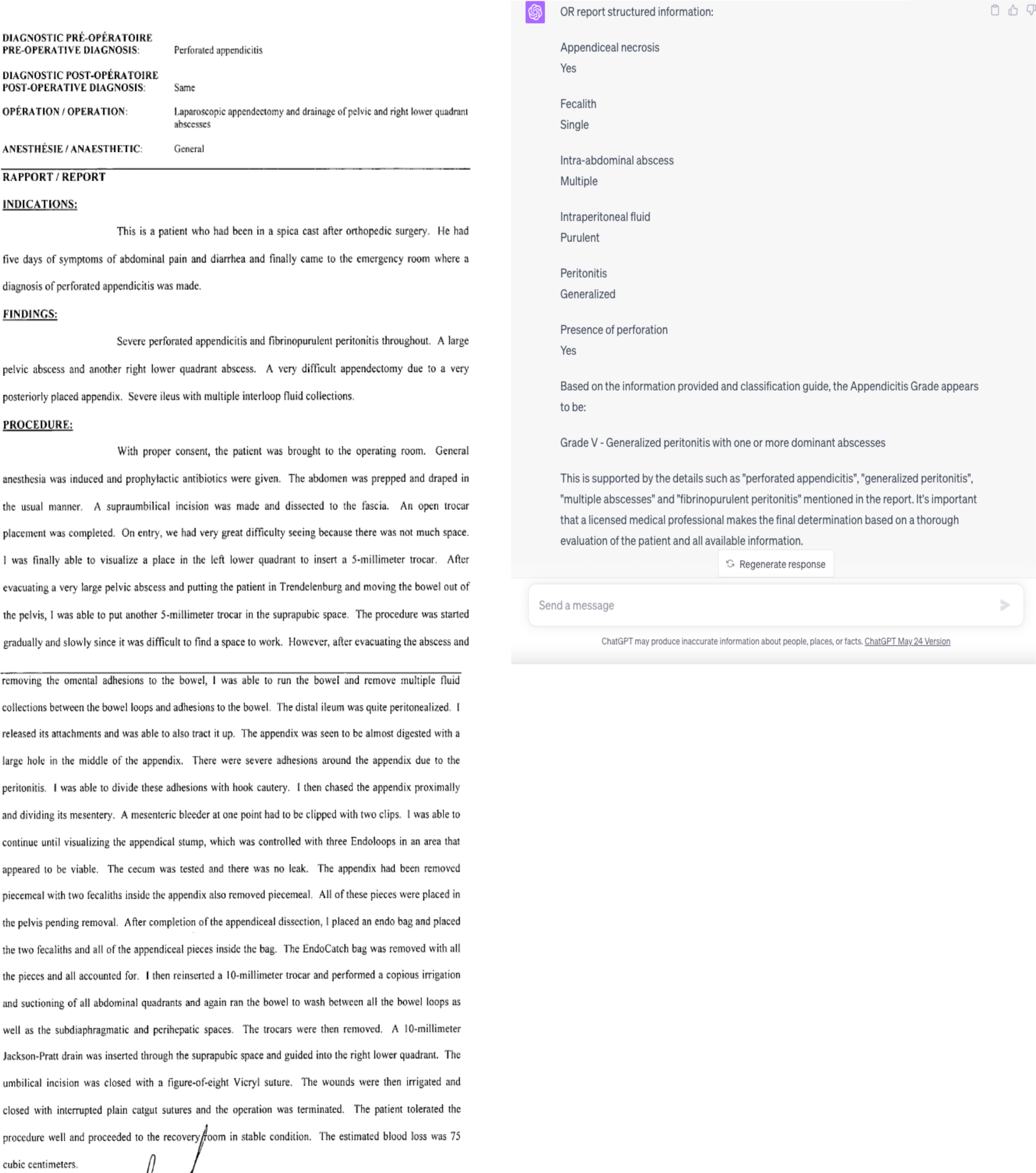
Operative Report & ChatGPT-4 Structured Data After Prompt.

**Fig. 3:**
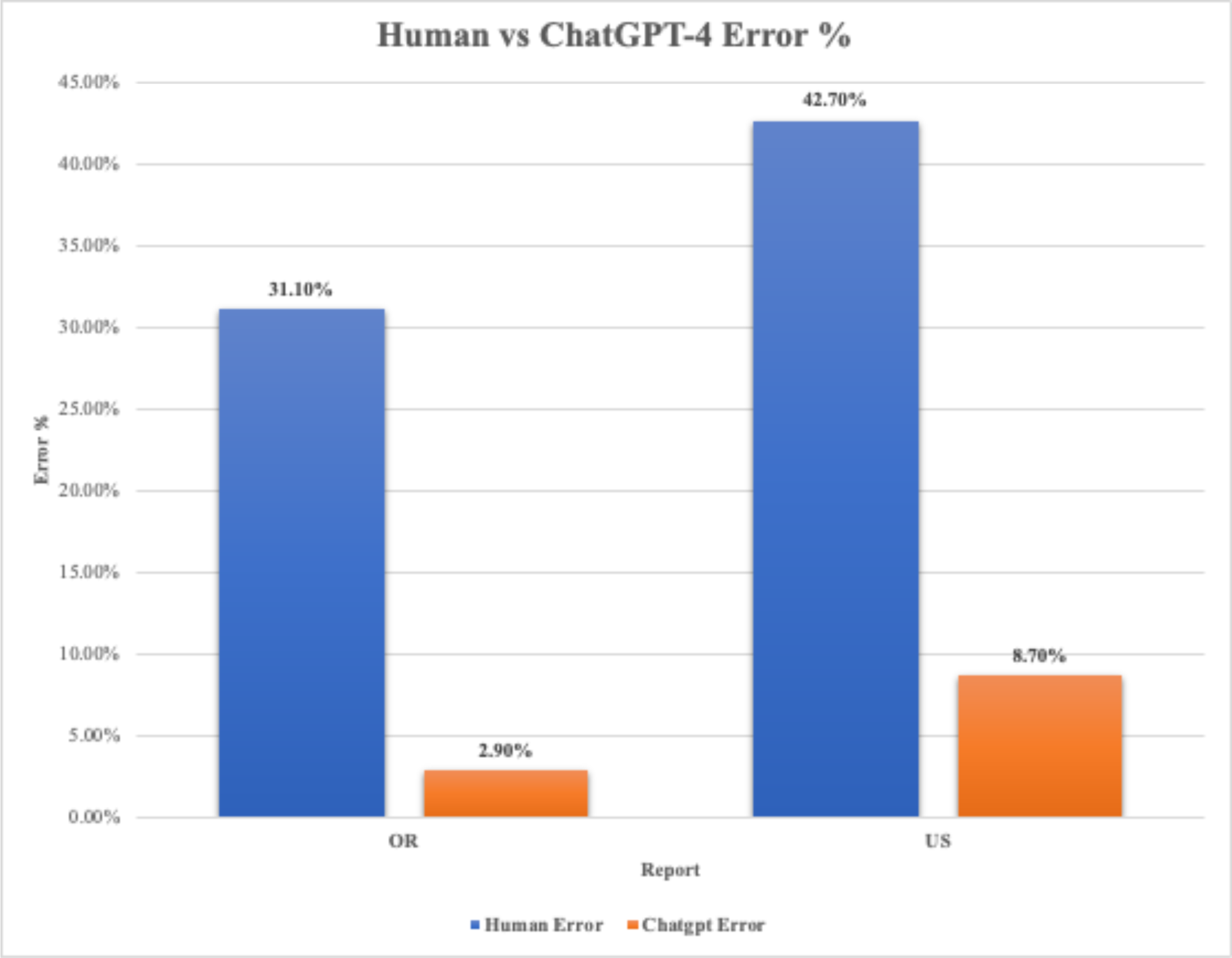
Human And ChatGPT-4 Errors In Data Extraction.

### Time required

We compared the time saved using the AI chatbot for data extraction vs human data extractors. The latter required an average of 20 minutes to extract data from patient records into the secure web-based software (REDCap). For the 103 records, this totaled 33.3 hours. By comparison, when using ChatGT-4, taking into account copying and pasting the required information from the reports, alongside the prompts, into the chatbox, and generating the required output from ChatGPT-4, task completion for one record required on average 30 seconds - totalling therefore 50 minutes for 103 records. The AI chatbot was therefore almost 40 times faster than the human extractors in this extraction/summarization task. It is however worth noting that currently ChatGPT-4 is limited to only allowing 25 entries every 3 hours. One other limitation is the occasional need to refresh the prompt thread, as the chatbot seemed to get “fatigued” and not respect exactly the constraints listed in the prompt after approx. ten records.

## Discussion

In this study we compared accuracy of data extracted by human data extractors, on detailed preoperative and operative data from the EMR of children operated for acute appendicitis, to ChatGPT-4 extraction data after specific prompts. The data was extracted from US and operative reports to estimate the PAG. After comparison between human data extractors and ChatGPT-4t to identify discrepancies, the latter was found to be more accurate in extracting data from both US and operative reports by 41.7% and 47.6%, respectively. Not only did it provide more accurate data extraction, but it made the process of data extraction 40 times faster.

To our knowledge, there are no other studies that directly compared free-text data extraction from the EMR between AI chatbots and human data extractors. Adamson et al [27] applied NLP to train, validate, and test the extraction of information from unstructured documents (e.g., clinician notes, radiology reports, lab reports, etc.) to output a set of structured variables required for real-world data analysis. The authors concluded that NLP enabled the extraction of retrospective clinical data from the EMR faster and more efficiently [27]. The authors did not however directly compare their model to human data extractors - their purpose was to show success in building an AI model that extracts accurate information from the EMR.

There are, however, several studies successfully documenting the use of AI chatbots in clinical settings to extract EMR information. Clinical text differs significantly from typical text used in general NLP, both in syntax and vocabulary [28]. As a result, the clinical NLP community often trains domain-specific models on clinical corpora, using language modeling strategies from the broader NLP community. However, in several such applications the performance gains were marginal compared to classical methods such as logistic regression [29,30]. Early studies using LLMs such as GPT-3 failed to show competitive results on biomedical NLP tasks [31,32]. One study developed a generic predictive model that covers observed medical conditions and medication uses [33]. This temporal model using recurrent neural networks was developed and applied to longitudinal time stamped EMR data. Encounter records (e.g. diagnosis codes, medication codes or procedure codes) were input into recurrent neural networks to predict diagnosis and medication categories for subsequent visits, showing improved accuracy compared to several baselines that are based on experts’ intuition about the dynamics of events in clinical settings [33]. Other groups have been able to predict the risk of 30-day readmission [34] and diagnose rare diseases [35] based on structured EMR data.

LLMs can also be used to generate accurate, relevant, comprehensive, and coherent answers to clinical questions based on hospital admission notes. One study demonstrated the accuracy, relevance, comprehensiveness, and coherence of the answers generated by AI chatbots (ChatGPT 3.5 and Claude) on a set of patient-specific questions [36]. Their results suggest that LLMs are a promising tool for patient-specific inquiries from clinical notes [36].

ChatGPT’s versatile nature and advanced NLP capabilities have made it a valuable tool across various domains, specifically in healthcare, education and scientific research. Some of the potential for ChatGPT use in healthcare include: (i) chatbots that can assist with patient triage, helping healthcare providers determine the urgency of a patient’s condition and the appropriate course of action [37]; (ii) medical diagnosis and treatment recommendations by analyzing patient data and symptoms [38], (iii) patient engagement and adherence - by providing personalized recommendations and reminders, helping patients stay on track with their treatment [39], (iv) clinical research and development through analysis of large amounts of clinical data, identifying patterns and trends that can be used to develop new treatments and interventions [40]. In the medical education and training sector: (i) AI has been beneficial in personalized learning by providing tailored recommendations in medical imaging to aid in diagnosing diseases such as surgical training and tutoring [41,42]. (ii) ChatGPT has the potential to assist with medical education, and potentially, clinical decision-making and knowledge development [43,44]. The application of ChatGPT in scientific research is multifaceted: (i) ChatGPT has been instrumental in transforming the way researchers interact with and interpret data [45]. (ii) ChatGPT is a powerful tool for hypothesis generation and testing, aiding researchers in conceiving new research questions and hypotheses[46]. Moreover, substantial progress has been achieved in discerning the extensive implications of ChatGPT in areas such as healthcare and education research, although these domains do present their unique challenges[38,47].

### Limitations

There are several limitations to the current study. Our data is specific to pediatric appendicitis, therefore the findings may not be generalizable to other types of medical records or patient populations - the performance of both the AI chatbot and the human extractors might differ when dealing with other types of data. However, with the proper prompts the chatbot should be able to successfully extract structured data from other datasets and populations. Another limitation was the natural presence of missing and/or incorrect data in the EMR, which could potentially impact the accuracy of both extraction methods. Such data input gaps and errors would however impact both extraction methods similarly. Moreover, the data set used in this study was relatively small. It is possible that the results would be different if a larger data set were used. Furthermore, it’s important to note that the accuracy evaluation was conducted by a single individual - a senior pediatric surgeon. While this could potentially introduce a higher risk of bias, the evaluator’s expertise in the field helps mitigate this concern.

AI chatbot performance is naturally very dependent on the LLMs and the content they have been trained on, in particular the size of the medical corpus. The other key limitation is the quality of the prompts used, which is related to the rapidly expanding expertise in the new field dubbed “prompt engineering” [48].

Both human and AI performance could have been affected by the quality and specificity of the pre-defined extraction criteria. The large number of errors identified after the manual data extraction was particularly concerning, raising doubts on the adequacy of the on-task content training and certification, despite satisfactory Kappa scores.

Finally, it’s worth noting that the use of any AI, such as AI chatbots, in analyzing and extracting patient data raises important ethical and privacy concerns. To address these concerns, only de-identified data was provided to ChatGPT. Despite these limitations, the results of this study suggest that AI chatbots have the potential to be a valuable tool in a healthcare research setting.

In spite of the valuable contributions made by AI in healthcare, it is important to acknowledge and address the challenges and ethical implications arising from its application. This includes a multitude of factors, such as: (i) model reliability and precision, wherein the AI could generate erroneous or deceptive information; (ii) Inherent bias in AI models can lead to biased and inequitable information and treatment suggestions, and the efficiency of ChatGPT may also be affected by dataset bias, where the quality and diversity of the data used for training could influence the model’s performance; (iii) overdependence on AI, consequently diminishing critical thinking and independent problem-solving skills; (iv) privacy issues due to chatbots’ access to extensive user data, triggering concerns over privacy and data protection; and (v) factual integrity, since LLMs can “hallucinate”, generating inconsistent or erroneous texts. It is thus crucial to verify the factual correctness of the generated content [47]. AI models also grapple with issues related to understanding context, ethical reasoning, conversational context, and generation of visual content. ChatGPT, while impressive, grapples with several challenges such as handling inappropriate requests, adjusting to user expertise levels, delivering personalized feedback, and dealing with multilingual queries and non-literal language. There are also ethical quandaries related to data privacy and security, intellectual property rights, transparency, accountability, and susceptibility to adversarial attacks. The potential influence of ChatGPT on human behavior prompts questions about personal autonomy. Bias and discrimination issues are also prevalent, as AI language models like ChatGPT are trained on extensive datasets which could inadvertently include biases, stereotypes, and prejudiced language. This could result in the model unknowingly generating offensive or harmful responses, thereby perpetuating these biases. Mitigating this problem necessitates refining the training data, enhancing the model’s structure, and implementing guidelines to ensure fairness and unbiased results[25,49,50]. Approaching these ethical considerations and challenges requires a forward-thinking attitude from developers, researchers, and the larger AI community. Collaborative efforts to recognize, comprehend, and resolve potential issues will ensure that AI language models like ChatGPT are developed and utilized in a responsible manner, maximizing their advantages while minimizing potential risks.

## Conclusion

Our study showed that ChatGPT outperformed manual data extraction for ultrasound and operative reports of acute appendicitis in accuracy, and correctly assigned the PAG score. Despite significant limitations in the clinical use of similar AI chatbots, these novel AI tools already show promise in rendering research tasks more accurate and significantly faster.

## Data Availability

All data produced in the present study are available upon reasonable request to the authors

