## Supplemental File for "Using Artificial Intelligence To Label Free-Text Operative And Ultrasound Reports For Grading Pediatric Appendicitis"

**Supplemental File A (1): Prompts Used for ChatGPT-4 Data Extraction**

**Ultrasound (US) Report**

**Ultrasound (US) Report PROMPT:**

*Please structure the following free-text ultrasound reports for pediatric appendicitis using only the headings and categories below:*

**US Report Template**

Ultrasound diagnosis

- Simple appendicitis
- Perforated appendicitis
- Normal appendix
- Appendix not visualized, fat stranding
- Appendix not visualized, no fat stranding
- Other

Appendix identified

- Yes
- Partial
- No
- Not recorded

Appendiceal maximum diameter in mm

Appendix location

- Medial (to IC valve)
- Retrocecal
- Other
- Not specified

Probe tenderness over appendix

- Yes
- No
- Not recorded

Appendix compressible

- Yes
- No
- Not recorded

Mesenteric fat stranding

- Yes
- No
- Not recorded

Fluid around appendix

Yes

No

Not recorded

Fluid in pelvis

Yes

No

Not recorded

Phlegmon or inflammatory mass

Yes

No

Not recorded

Bowel thickening

Yes

No

Not recorded

Presence of fecalith

Yes

No

Not recorded

Intra-abdominal abscess

None

Single

Multiple

Not mentioned

*The free-text reports will start now:*

**Supplemental File A (2): Prompts Used for ChatGPT-4 Data Extraction**

**Operative Report (OR)**

**Operative Report (OR) PROMPT:**

*Use the operative report (OR) template below to structure the information needed from any OR I will provide, and attempt to the best of your abilities to assign to each a perforation grade based on the classification below.*

**OR Template**

Appendiceal necrosis

 Yes

 No

 Not mentioned

Fecalith

None

 Single

 Multiple

 Not mentioned

Intra-abdominal abscess

None

 Single

 Multiple

 Not mentioned

Intraperitoneal fluid

 None

 Serous

 Seropurulent

 Purulent

 Not mentioned

Peritonitis

 None

 Localized

 Generalized

 Not mentioned

Presence of perforation

 Yes

 No

 Not mentioned

Appendicitis Grade

Grade 0 - Normal Appendix (May include terms such as “Normal” or “Unremarkable appendix” or “white appendix”)

Grade 1 - simple appendicitis (May include terms such as “appendix non-perforated” or “simple appendicitis” or “Appendix” or “ “inflamed” or “phlegmonous” or “gangrenous” or or “dilated” or “necrotic” or “injected” or “edematous” “non-compressible”)

Grade II - Localized or contained perforation (May include terms such as “Perforated appendicitis” or “Complex appendicitis” or “Appendix perforated” or “leaking pus” or “pus” or “fluid” or “collection around the appendix” or “Appendix sealed” or “covered by omentum”)

Grade III - Contained abscess with no generalized peritonitis (May include terms such as “Perforated appendicitis” or “Complex appendicitis” or “Single abscess” or “Break into” or “unroof a collection” or “pocket of pus” or “Interloop abscess”)

Grade IV - Generalized peritonitis with no dominant abscess (May include terms such as “Perforated appendicitis” or “Complex appendicitis” or “Generalized peritonitis” or “Pus in all/four quadrants” or “Diffuse peritonitis”or “Fibrinopurulent peritonitis”).

Grade V - Generalized peritonitis with one or more dominant abscesses (May include terms such as “Perforated appendicitis” or “Complex appendicitis” or “Single or multiple abscesses” or “Break into” or “unroof collection(s)” or “Pocket(s) of pus” or “Interloop abscess(es)” or “Pus in four quadrants with abscess” or “Diffuse peritonitis” or “Fibrinopurulent peritonitis”).

*The free-text reports will start now:*
